# Plaque Contact Surface Area and Flow Lumen Volume Predict Stroke Risk in Extracranial Carotid Artery Stenosis

**DOI:** 10.1101/2021.10.01.21264200

**Authors:** Ryan Gedney, Ethan Kung, Veena Mehta, Adam Brown, Matthew Bridges, Ravi Veeraswamy

## Abstract

The standard indication for intervention in asymptomatic disease is currently percent stenosis in the internal carotid artery as measured by the NASCET method, which remains limited in discriminating power. CT angiography (CTA) is widely used to calculate NASCET stenosis but also offers the opportunity to analyze carotid artery plaques from a morphological perspective that has not been widely utilized. We aim to improve stroke risk stratification of patients with carotid artery stenosis using plaque 3D modeling and image analysis. Patients with CTAs appropriate for 3D reconstruction were identified from an NIH designated stroke center database, and carotid arteries were segmented and analyzed using software algorithms to calculate contact surface area between the plaque and blood flow (CSA), and volume of the flow lumen within the region of the plaque (FLV). These novel parameters factor in the 3D morphometry inherent to each carotid plaque. A total of 134 carotid arteries were analyzed, 33 of which were associated with an ipsilateral stroke. Plaques associated with stroke demonstrated statistically significant increases in average CSA and FLV when compared to those not associated with stroke. When compared to NASCET percent stenosis, CSA and FLV both demonstrated a larger area under the receiver operating characteristics curve (AUC) in predicting stroke risk in patients with carotid stenosis. The data presented here demonstrate morphological features of carotid plaques that are independent of NASCET criteria stratification and may present an improved method in assessing stroke risk in patients with carotid artery stenosis.

## Introduction

Stroke accounts for 1 in 19 deaths and is the 5th leading cause of mortality in the United States. Over 795,000 people per year suffer from stroke and 150,000 people per year die from strokes in the U.S.^1–3^ Beyond mortality, the morbidity caused by strokes can be devastating. Between 2-3% of the total population in the US claim long-term disability in employment secondary to stroke.^4^ Approximately 20% of ischemic strokes are caused by extracranial carotid artery stenosis,^2,3,5^ and the 5-year stroke or death risk is upwards of 11% in asymptomatic patients with extracranial disease receiving medical therapy alone.^5^ Studies have a shown that the 5-year stroke or death risk in asymptomatic patients can be reduced from 11% to 5% with appropriate carotid intervention.^5^ There is a narrow therapeutic window however and for patients without symptoms, 22 carotid interventions will have to be performed to prevent 1 stroke.^6^

In patients with carotid artery stenosis, the goal is to identify lesions that are predisposed to causing a cerebral stroke. The inability to identify high-risk lesions has been a hurdle to implementing a more effective strategy for intervention leading to persistently high stroke rates and, simultaneously, possibly unnecessary interventions. The current indication for intervention in asymptomatic carotid stenosis is ‘percent stenosis’ in the internal carotid artery as measured by the NASCET method.^7^ This is a calculation of the narrowest diameter of the internal carotid artery compared to the presumed normal caliber of the distal internal carotid artery. This only accounts for lumen diameter within two planes (Fig 1), and there is no consideration given for the morphology of the lesion, length of the involved segment or patient demographics. As demonstrated in Fig 1, two clearly morphologically different carotid artery plaques can be stratified as identical by NASCET. A plaque with tortuous surface morphology and longer length should present a much higher risk profile than a short smooth plaque, even if the percent stenosis is identical.

**Figure 1:**
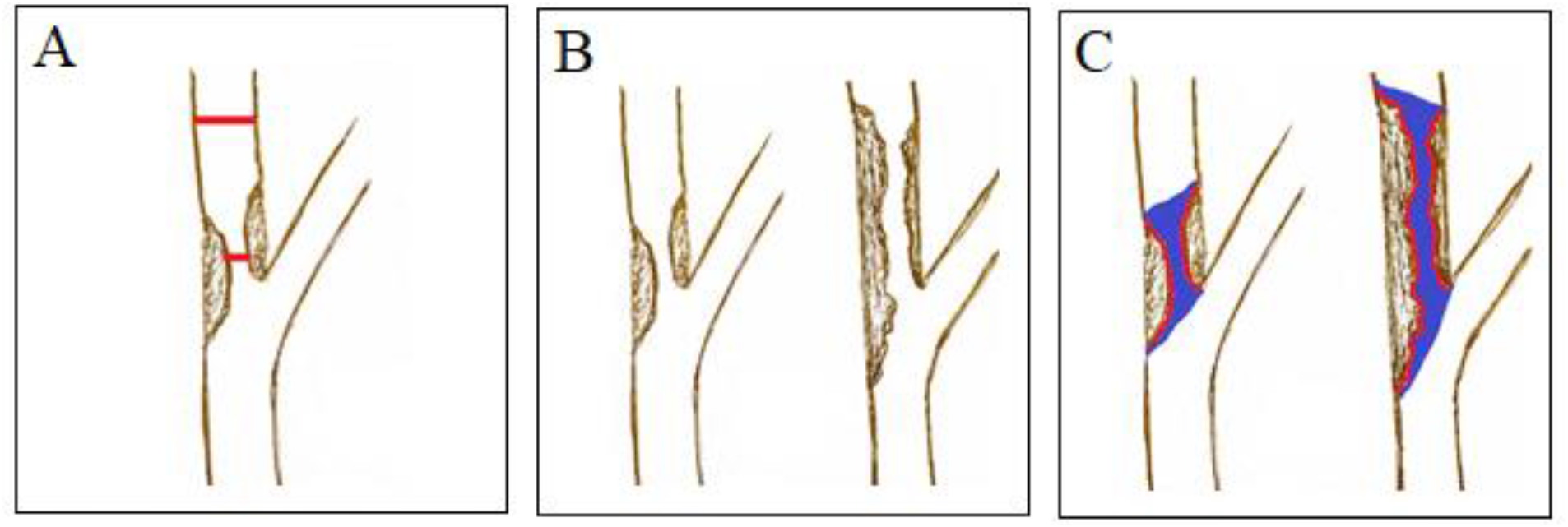
Visual representation of (A) the planar analysis of diameter that is utilized when measuring NASCET percent stenosis via CT angiography, (B) two carotid artery plaques that are measured as identical based on the NASCET method, but clearly demonstrate significantly different morphologies that intuitively should affect their risk stratification, and (C) contact surface area (outlined in red), and flow lumen volume (highlighted in blue).

Previous attempts to improve on the NASCET methodology have been underwhelming.^8–11^ The intense study on plaque characterization and attempts to identify vulnerable lesions have not led to clinical improvements in risk stratification. The use of magnetic resonance imaging to characterize plaques is challenging to incorporate into an efficient workflow. A large, randomized trial comparing intervention to medical management (CREST-2) is underway, but it is not designed to answer the question of how best to determine the risk of a given lesion^12^. There is a clear need for improved, reliable risk stratification of carotid lesions which will allow for targeted intervention and a reduction of overall stroke incidence.

We have identified two novel variables, contact surface area (CSA) and flow lumen volume (FLV), that provide a more accurate assessment of stroke risk and are easily derived from conventional CT scans of carotid arteries obtained as standard-of-care. The contact surface area (CSA) is a variable we have defined as the area over which flowing blood is in contact with plaque within the lumen of the artery, outlined in red in Fig 1, and flow lumen volume (FLV), is the volume of the column of blood within the region of plaque, highlighted in blue in Fig 1. Plaque irregularities and size have already been demonstrated to be associated with stroke,^13,14^ and contact surface area and flow lumen volume offer a metric to quantify this property. Intuitively, greater contact surface area offers greater opportunity for flowing blood to affect shear stresses on a vulnerable plaque, instigating thrombotic or embolic events. Similarly, FLV allows us to capture information about the plaque that lumen diameter alone cannot, including plaque contours and length.

We aim to test these variables, CSA and FLV, using 3D modeling and analysis for assessing stroke risk in patients with carotid stenosis, and hypothesize that these markers can do so with a higher sensitivity and specificity than current methods.

## Methods

A single center, HIPAA compliant database of patients from The Medical University of South Carolina stroke center, an NIH designated stroke center, was assembled. Institutional approval was granted via Institutional Review Board. Within the database, strokes adjudicated to have been caused by embolization from the ipsilateral carotid artery were identified. Patient demographic data and the corresponding carotid CTAs were downloaded into a secured server. We also downloaded patient data and carotid CTAs not associated with a stroke.

CTA scans for each patient were reviewed, and those appropriate for 3-D reconstruction were downloaded on a secure, encrypted server at the site of the stroke center, and then stored in an institutional approved, password protected, cloud-based storage system. Scans were deemed appropriate for 3-D reconstruction based on several criteria: sufficient contrast between carotid artery, plaque, and surrounding structures., free of any artifact, including metal implants, that interferes with appropriate contrasting and visualization of the carotid artery and plaque, CTA slices ≤1mm. Downloaded CTA scans were de-identified at the time of downloading and stored in a coded format as to be able to identify the appropriate associated patient with each stored scan. The percent stenosis listed in the radiologist report for each CTA was recorded for each internal carotid artery with demonstrated stenosis.

CTA scans were each uploaded to 3D Slicer [version 4.11.20200930, BWH and 3D Slicer Contributors], an open source 3-D segmentation and analysis software (Fig 2). This software allows for scans to be uploaded accounting for slice thickness and dimensions for appropriate 3D modeling and calculations based on 3D models. Thresholding was used to highlight Hounsfield units equivalent to that of contrast in each CTA, and manual cropping tools built into the software were then utilized to isolate the region of the internal carotid artery associated with atherosclerotic plaque (Fig 2). Arteries that did not have associated plaque in the internal carotid artery or had 100% occlusion were excluded. Once the appropriate Hounsfield unit identification and manual cropping had been performed, the region was segmented, generating a 3-D model of the internal lumen of the internal carotid artery only within the region of plaque (Fig 2). This 3-D model can be understood as a mold of the internal surface of the carotid artery plaque and containing the volume of blood within the region of the plaque. This allowed for excellent visualization of plaque contours as seen in Fig 2-D. Internal software algorithms within the 3D Slicer environment were then used to calculate the surface area and volume in units of mm^2^ and mm^3^, respectively. These values were recorded as contact surface area and flow lumen volume for each plaque in association with their respective patient.

**Figure 2:**
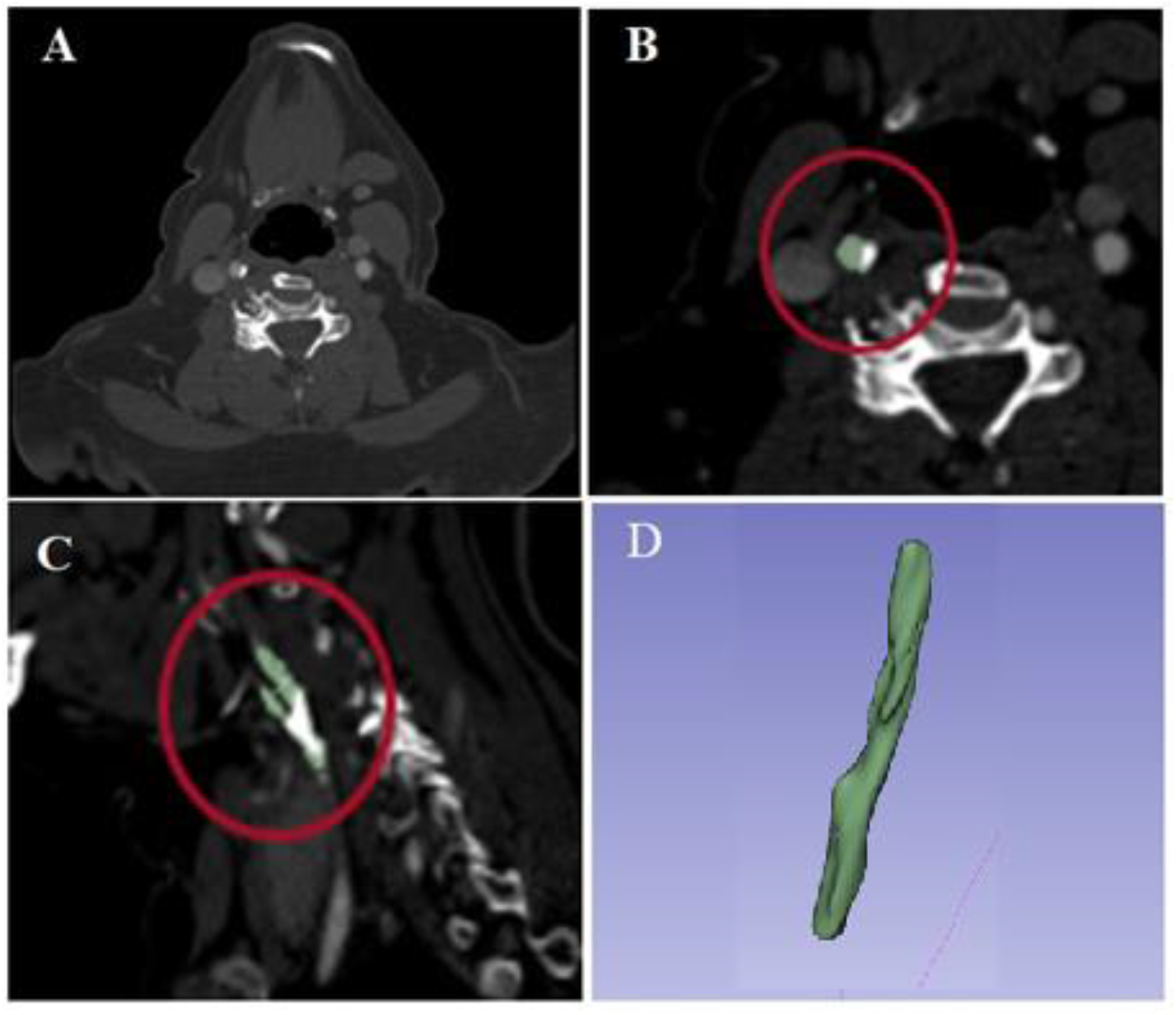
(A) Representation of appropriate windowing of CTA for 3D modeling with 3D slicer, (B) and (C) Hounsfield unit boundary identification utilized to isolate the carotid artery from the rest of the CTA, (D) cropped segmented 3D model of the internal surface of the carotid bifurcation

**Figure 3:**
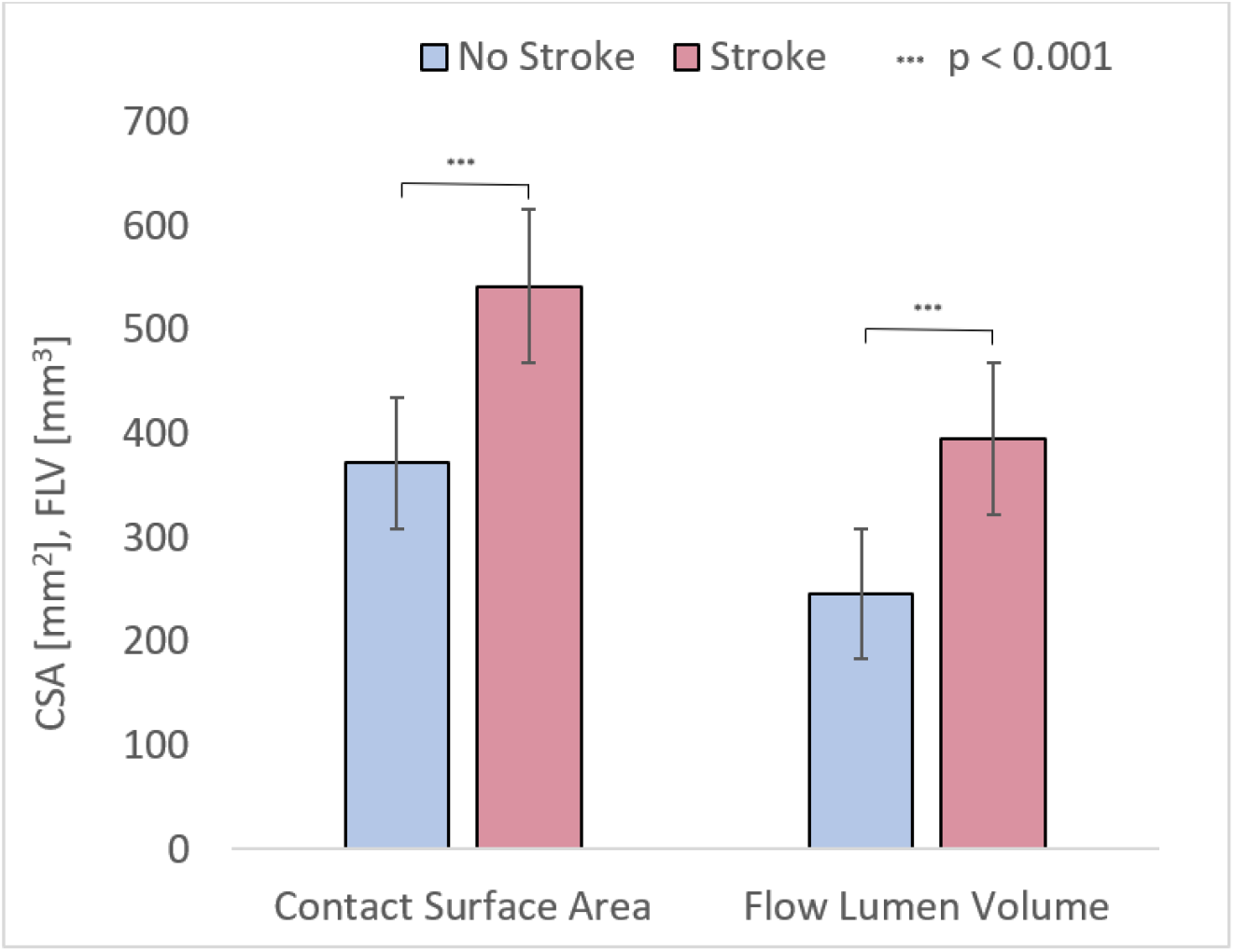
Both CSA and FLV demonstrate statistically significant increases in plaques associated with stroke.

**Figure 4:**
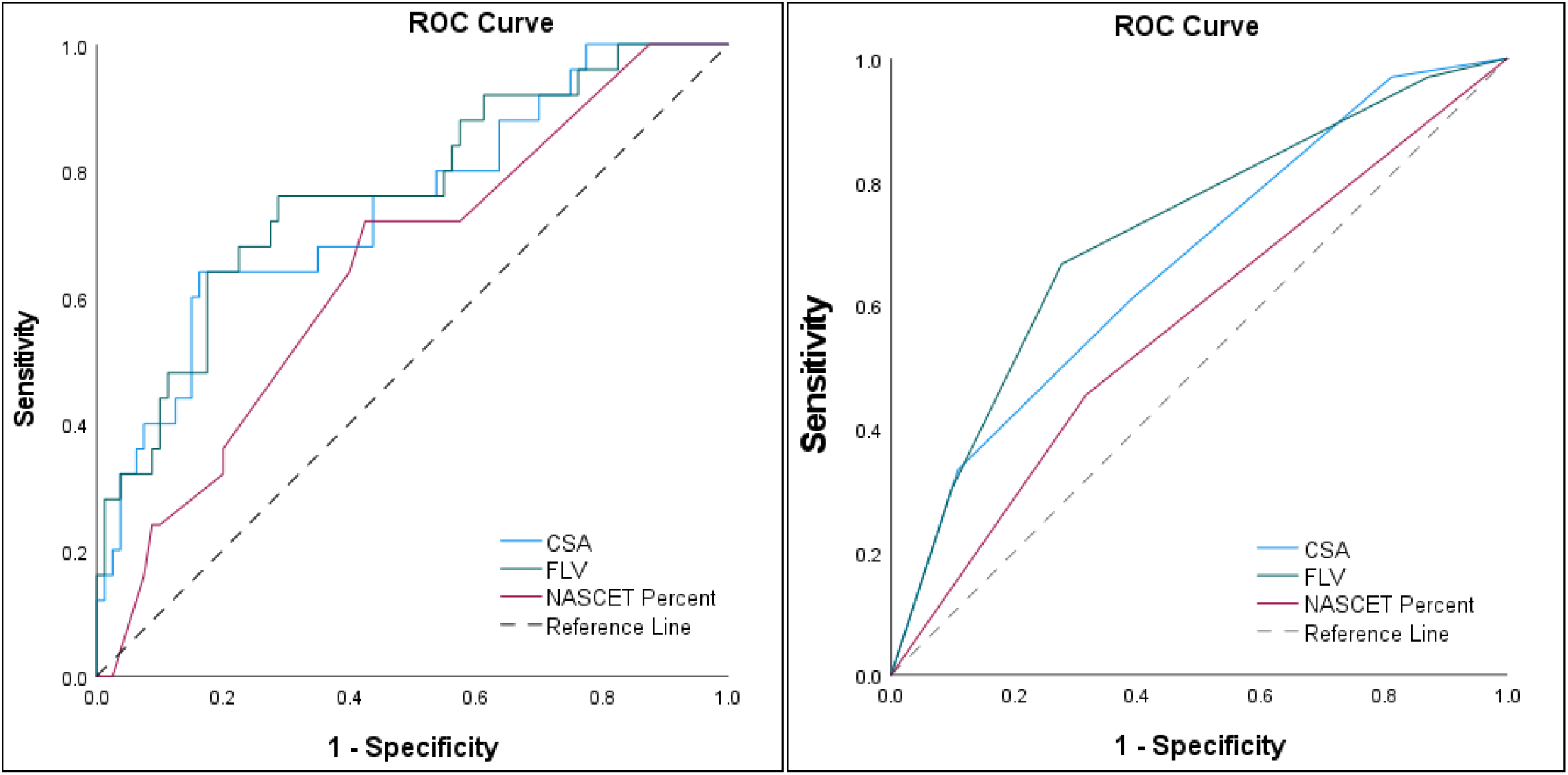
ROC curve built with CSA, FLV, and NASCET percent stenosis as continuous variables (left) and categorical variables (right). As continuous variables, CSA and FLV demonstrated AUC of 0.742 and 0.766, respectively, and NASCET percent stenosis demonstrated an AUC of 0.646. As categorical variables, NASCET percent stenosis was categorized according to standard <50%, 50-70%, and >70% percent stenosis groups. CSA was categorized as follows in units of mm^2^: <203, 203-413, 413-623, and >623. FLV was categorized as follows in units of mm^3^: <115, 115-283, 283-450, and >450. CSA and FLV demonstrated AUC of 0.669 and 0.714, respectively, and NASCET percent stenosis demonstrated an AUC of 0.569.

Regression analysis accounting for demographics, comorbidities, and medication use was utilized to statistically analyze the contact surface area and flow lumen volume independently in their association with stroke and non-stroke patients. Regression analysis was also done for NASCET percent stenosis of each carotid plaque analyzed. Receiver operator curves were generated in comparing continuous variables CSA, FLV, and NASCET percent stenosis as predictors of stroke risk in the carotid arteries analyzed. A receiver operator curve was also generated for CSA, FLV, and NASCET percent stenosis as categorical variables. NASCET percent stenosis was stratified into categorical variables <50%, 50-70%, and >75%, and CSA and FLV were stratified into categorical variables based on standard deviations from the mean.

## Results

A total of 134 internal carotid arteries were analyzed with 3-D simulation, 33 of which were associated with an ipsilateral stroke, and 101 of which were not. Plaques associated with stroke demonstrated an increased average CSA of 541.52 mm^2^ when compared to those not associated with stroke which had an average CSA of 371.18 mm^2^ [p < 0.001]. Plaques associated with stroke demonstrated increased average flow volume of 394.64 mm^3^ when compared to plaques not associated with stroke which had an average FLV of only 245.92 mm^3^ [p < 0.0001]. Plaques associated with stroke demonstrated increased average NASCET percent stenosis of 69% when compared to plaques not associated with stroke which had an average NASCET percent stenosis of 58% [p = 0.01].

Compared to NASCET percent stenosis, CSA and FLV demonstrated increased area under the curve of 0.74 and 0.77, respectively. Percent stenosis as a continuous variable demonstrated an AUC of only 0.65. This relationship held when categorizing CSA and FLV based on standard deviations from the mean and comparing to percent stenosis as risk categories. When comparing categorically, area under the curve for CSA (0.67) and FLV (0.71) remained higher than NASCET percent stenosis (0.57). NASCET percent stenosis was categorized <50%, 50-70%, and >70%.

## Discussion

Risk stratification for extracranial carotid bifurcation lesions has remained surprisingly static since the publication of the NASCET and ACAS trials. Despite advances in understanding atherosclerosis, plaque instability and fluid mechanics, determining which carotid lesions warrant intervention still hinges upon a simple 2-dimensional ‘percent stenosis’ in the internal carotid artery. The simplicity of this calculation ignores fluid mechanics inherent to a complex 3-dimensional lesion. Carotid plaques with greater surface area are associated with increased surface roughness, considered as a material property, because of the inevitable bumps and curves that line the contact surface. Considering the standard Moody Diagram^15^ and using a Reynold’s number of roughly 2000 for blood, increasing roughness results in flow that is more likely to be turbulent flow. Turbulent flow is directly related to flow separation that has been demonstrated to contribute to development and progression of atherosclerotic plaque.^16^ These changes in fluid dynamics directly affect cell signaling, further contributing to inflammation and an environment for plaque development, rupture, and distal particulate embolization.^17^ Despite advances in computational capabilities, image acquisition and fluid dynamics, little work has been done to measure the relevance of carotid plaque morphology in a clinically applicable manner. Our study focuses on measuring the effect of the shape and length of the plaque utilizing easily accessible CTA scans of the carotid artery. The CSA and FLV are both a measure of the dynamic surface, contours and length of the plaque and provide a more nuanced measure of the instability and embolic risk of the plaque than percent stenosis alone. The improved sensitivity and specificity of these methods compared to the NASCET method is encouraging. The relatively poor correlation between NASCET percent stenosis and stroke in our ‘real world’ dataset was surprising. Together, these data underscore the need for a more refined approach to risk stratification for extracranial disease.

It is also important to note that improvements upon existing methodology must be compatible with clinical integration. The use of MRI is simply not feasible for most patients. It is limited by patient access, time, and cost. While the flow data and plaque characterization from MRI/MRA are informative, it remains largely untenable as a tool for routine patient care. Carotid doppler examination has the converse problem – it is easily available, but the operator dependent nature of results and inability to validate anatomic criteria from carotid ultrasounds has limited the enthusiasm for expanding the data derived from carotid ultrasounds beyond the currently used flow metrics.^18^

Our work seeks to fully exploit all the data available from CTA that are currently routinely used in clinical care. A limitation of our study is that we analyzed CTA obtained at the time of the stroke. While there is no evidence that the gross morphology of a carotid bifurcation lesion changes at the time of embolization, it would be ideal to have a CTA prior to the stroke itself. Even in a large data repository such as ours, we found only 6 patients with a CTA within a year of their cerebral event – clearly not enough to interpret changes meaningfully. Further work will aim to expand our dataset to other centers such that we can accrue enough CT studies of patients within 12 months of their cerebral event to definitively address this limitation. Further study with a prospective trial will ultimately be needed and seems reasonable to undertake with these results. We simultaneously present two distinct parameters, CSA and FLV, as potential candidates to supplant stenosis as stroke predictors. We are not able to distinguish one of these as superior and larger studies will be needed to understand the nuances between the two. At present, calculating CSA and FLV is a manual process requiring some level of sophistication with 3-D image manipulation. Our group was able to teach this to novice students in a few weeks’ time, and we are currently working on automating the process completely.

In addition to expanding the database, future directions will incorporate patient demographics, computational fluid dynamics and utilize machine learning to further improve the accuracy of predictive modeling.

## Conclusion

The data presented here validates the novel variables CSA and FLV, as they quantify morphological features of carotid artery plaques and demonstrate association with stroke. Increased contact surface area and flow lumen volume are associated with stroke and may present an improved protocol in assessing stroke risk in patients with carotid artery stenosis.

## Data Availability

Data is available per request.

---

**Table 1.**

| **Demographics and Comorbidities** |  |  |  |
| --- | --- | --- | --- |
|  | **Stroke Group [%]** | **No Stroke Group [%]** | **p-value** |
| **Total Number (CTAs)** | 42 | 51 | - |
| **Age** | 65.7 | 68.5 | 0.07 |
| **Male Gender** | 55 | 59 | 0.7 |
| **CAD** | 36 | 41 | 0.6 |
| **DM** | 26 | 27 | 0.89 |
| **HLD** | 67 | 84 | 0.05 |
| **HTN** | 88 | 78 | 0.22 |
| **TIA** | 19 | 10 | 0.21 |
| **Atrial Fibrillation** | 17 | 12 | 0.5 |
| **Smoking** | 79 | 80 | 0.83 |
| **Obesity** | 24 | 45 | 0.03* |
| **PVD** | 17 | 24 | 0.42 |
| **HF** | 10 | 14 | 0.54 |
| **CKD** | 7 | 12 | 0.46 |
| **ASA** | 57 | 71 | 0.18 |
| **Statin** | 83 | 92 | 0.19 |
| **Plavix** | 40 | 35 | 0.61 |

**Table 2.**

| **Regression** |  |  |  |
| --- | --- | --- | --- |
|  | **No Stroke** | **Stroke** | **p-value** |
| **Contact Surface Area [mm2]** | 371.18 | 541.52 | <0.001 |
| **Flow Lumen Volume [mm3]** | 245.92 | 394.64 | <0.001 |
| **NASCET Percent Stenosis [%]** | 58 | 69 | 0.01 |

